# Dried blood spots analysis for targeted and non-targeted exposomics

**DOI:** 10.1101/2025.04.12.25325612

**Authors:** Vinicius Verri Hernandes, Maximilian Zeyda, Lukas Wisgrill, Benedikt Warth

## Abstract

Dried blood spots (DBS) are an established sample type, widely used in newborn screening programs for monitoring metabolic diseases. Their minimally invasive nature offers great promise for assessing chemical exposures, particularly during early life stages and in large-scale epidemiological studies. However, comprehensive evaluations of key analytical parameters such as extraction efficiency and matrix effects across multiple chemical classes remain limited. Moreover, the promising approach of broadly combining targeted and non-targeted mass spectrometric data evaluation remains unexplored in DBS small-molecule omics. Here, we present an optimized LC-HRMS workflow for combined exposomic and metabolomic analysis in DBS samples. Four extraction protocols were systematically compared, with analytical performance evaluated for >200 structurally diverse toxicants, pollutants, and other key biomarkers. The optimized protocol demonstrated acceptable recoveries (60–140%) and reproducibility (median RSD: 18%) for a majority of compounds. Matrix effects showed a median value of 76% (median RSD: 14%). In a proof-of-principle study, twelve exposure compounds of the target panel with diverse physicochemical properties were identified in real-life samples, with several reported for the first time in DBS human biomonitoring. Complementary non-targeted analysis further expanded the detectable chemical space, enabling reliable annotation of additional exposures. Moreover, high-confidence identification of endogenous metabolites, including amino acids, biogenic amines, fatty acids and acylcarnitines demonstrated the capacity to capture a broad snapshot of the human metabolome. These findings support the use of DBS for integrated exposomics and metabolomics applications, providing toxicological and biological insights from low-volume samples in both, prospective and retrospective studies.

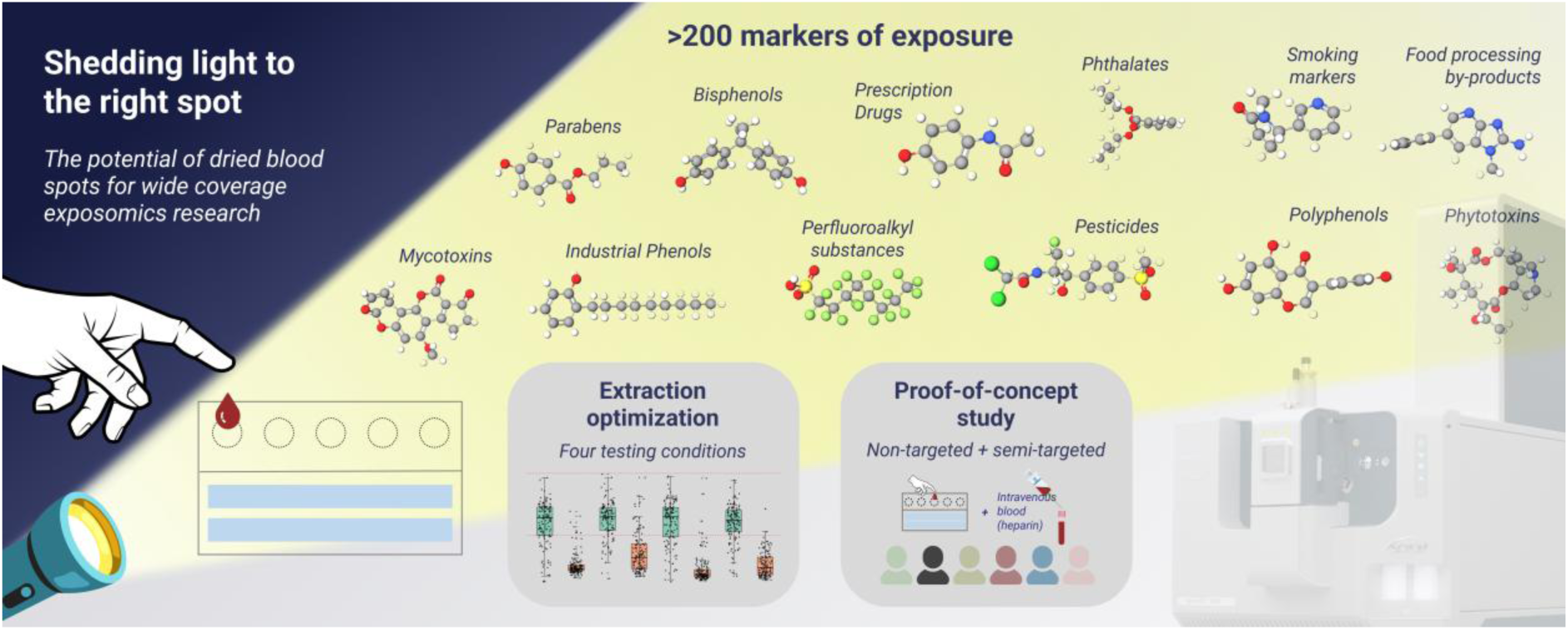

## 1. Introduction

Exposomics is a recent concept first introduced by Wild^1^ 20 years ago and further refined since then^2–4^ to the current understand of *the integrated compilation of all physical, chemical, biological, and psychosocial influences that impact biology*^5^. As such, the science of exposomics can be defined as the field which studies how the exposome can impact biological systems by integrating discovery-based analysis of environmental influences on health^5^. Given this prospect and the inherent variability of environmental exposures, the complexity of measuring the exposome throughout our life-span becomes evident.

To partially overcome this challenge, getting “snapshots” from key elements of the chemical exposome, particularly during susceptible stages of life, pose a practical approach^6^. Current efforts frequently focus on characterizing the exposome at sensitive/critical stages of life, such as pregnancy, early-life and reproductive years, among others^78^. There is vast evidence supporting the Developmental Origins of Health and Disease (DOHaD)^9,10^, which links periconceptual, fetal and early infant phases of life with long-term development^11^ and disease, including neurodegenerative diseases, breast cancer, cardiovascular disease, metabolic syndrome, infertility and asthma, among others^12–14^. Nevertheless, there is still an underlying gap regarding prenatal and infants exposure to xenobiotic mixtures, related to factors such as the lack of methodological approaches for assessing a broad range of xenobiotics, the scarce data on co-exposure for allowing holistic risk assessment and the focus towards large-scale (epidemiological) studies^8,15^.

Considering the need for comprehensive yet cost-efficient early-life exposure assessment at an epidemiological level, newborn screening programs (NBS) seem to be a promising fit-for-purpose option based on its inherent large-scale scope. These programs screens newborn within the first days of life for up to 70 distinct diseases that can be treated after early diagnosis^16^. Established in the early 1960s, the programmes developed from the pioneering work of Robert Guthrie and Ada Susi on the diagnosis of phenylketonuria and are now widely established with 39 million babies, roughly 28% of all newborns, screened in 2023^17,18^. NBS programs rely on the collection of a minimally invasive sample type, namely dried blood spots (DBS), for which a blood droplet obtained from the newborn heel is deposited on a paper card (named Guthrie cards) and analysed mostly via tandem mass spectrometry (MS/MS)^17^. Typically, residual DBS samples are stored, resulting in extremely valuable biospecimen collections available globally, for instance in Sweden (archiving residual DBS since 1975)^17^, Denmark (archiving DBS since 1982)^19^, Argentina (4.6 mi residual DBS stored over 25 years)^20^ and the USA, where 45% of the total newborn population stored for more than 21 years^21^.

Therefore, archived DBS are a highly valuable yet underutilized source for early-life exposomics research^22^. This includes retrospective analysis, being able to provide a chronological and comprehensive overview of exposure patterns from an epidemiological perspective. Recent works have reviewed the advancements in the human biomonitoring (HBM) of chemical exposures based on DBS samples^16,19,23–25^. Comparably extensive research was performed on volatile organic compounds (e.g. PCBs and PDBEs)^26–28^ and inorganic contaminants^29^ such as lead^30–33^, cadmium^30,33^ and mercury^30,33–35^. On the contrary, there is still very limited chemical space coverage for LC-MS-based methodologies, with a strong focus on targeted approaches for cotinine^36^, BPA^37^ and PFAS^37^ in the published literature. Additional specific chemical classes include the analysis of parabens^38^, mycotoxins^39^ and acrylamide^40^. To overcome limited chemical space coverage, suspect screening and non-targeted analysis (SSA and NTA, respectively) are emerging as promising tools. Nevertheless, whole blood, urine, placental tissue, breast milk, teeth and amniotic fluid are currently more established matrices when focusing on NTA and SSA studies for early-life exposure as recently reviewed by Guerrini *et al*^41^. Considering the contradiction between the great potential and the under usage of DBS for exposome research in an early-life context, it becomes essential to understand the real capabilities and limitations arising from this complex matrix.

Here, we tested four different solvent mixtures for a highly diverse set of >200 xenobiotics in DBS for a previously described LC-HRMS workflow integrating exposomics and metabolomics^42^. The first systematic evaluation of extraction recovery, electrospray-related matrix effects and the limits of detection for a semi-targeted evaluation of >200 organic environmental toxicants, including bisphenols, PFAS chemicals, mycotoxins, parabens, phthalates and others is provided, highlighting the potential and limitations of such an approach. Subsequently, exposure patterns and metabolic profiling were obtained from a proof-of-principle experiment with real-life samples from healthy adult donors, demonstrating the capacity of NTA analysis to enrich chemical diversity for both exogenous and endogenous compounds. This benchmarking data represents a critical step in laying an omic-scale foundation for next-generation HBM (targeted approach) and non-targeted exposomics and metabolomics (discovery-based approach) research utilizing DBS.

## 2. Material and Methods

### 2.1. Chemicals

Solvents used in all steps including mainly sample extraction and LC-MS analysis were LC-MS grade. Analytical standards for 223 xenobiotics were used to prepare a multicomponent standard solution at seven concentration levels. The selection of compounds intended to represent a highly diverse exposure panel for a high number of chemical classes, including phthalates, perfluoroalkyls substances (PFAS), parabens, polyphenols, mycotoxins, phytoestrogens, among others. Detailed information on reference standards including concentrations and spiking levels are provided in **Table S1.** Concentrations were compound-dependent and optimized in previous works^43–45^ based on exposure levels as well as instrument sensitivity. For most compounds, concentrations ranged from low to high ng/mL. Levels 0.8 and 20 Table S1) were used in spiking experiments and are referenced as levels ‘low’ and ‘high’, respectively.

Labeled Bisphenol A (Bisphenol-A-diphenyl-^13^C_12_), was purchased from Sigma-Aldrich (Catalogue number: 720186, Lot number: MBBB1923B). Labeled azithromycin (Azithromycin-d3) was purchased from TRC (Catalogue number: A927002). A mixture of labelled PFOS and PFOA (PFOS/PFOA extraction standard mixture, 13C8, 99% at 2000 ng/mL) was purchased from Cambridge isotope Laboratories (Catalogue number: ES-5571, Lot number: PR-28266). All chemicals were stored at –20°C until use.

### 2.2. Blood and dried blood spot collection

Whole blood was collected from six donors (three males, three females) in heparin tubes. Additionally, five dried blood spot samples were produced for each participant via fingertip collection of the ring finger after carefully cleaning with LC-MS grade isopropanol. The collection was performed by each participant to mimic self-collection and real-life sample quality using a Whatman 903 Protein Saver card. Participants were instructed to prioritize the falling-drop collection over hanging-drop collection whenever possible, and to avoid fingertip contact with the collection card as much as possible. A blood aliquot was used to determine haematocrit level and additional biochemical parameters using a Radiometer ABL90 Flex Plus blood gas analyser. Detailed information on biochemical parameters as well as additional participant metadata can be found in **Table S2.** The consent to participate in the study was obtained by the volunteers prior to study inclusion and the study approved by the ethics committee of the Medical University of Vienna (application number 1716/2023). The samples were stored at room temperature for six months, the total time needed to run and evaluate the comparison on the different solvent mixtures, further described in section 2.5 and Figure 1.

**Figure 1.**
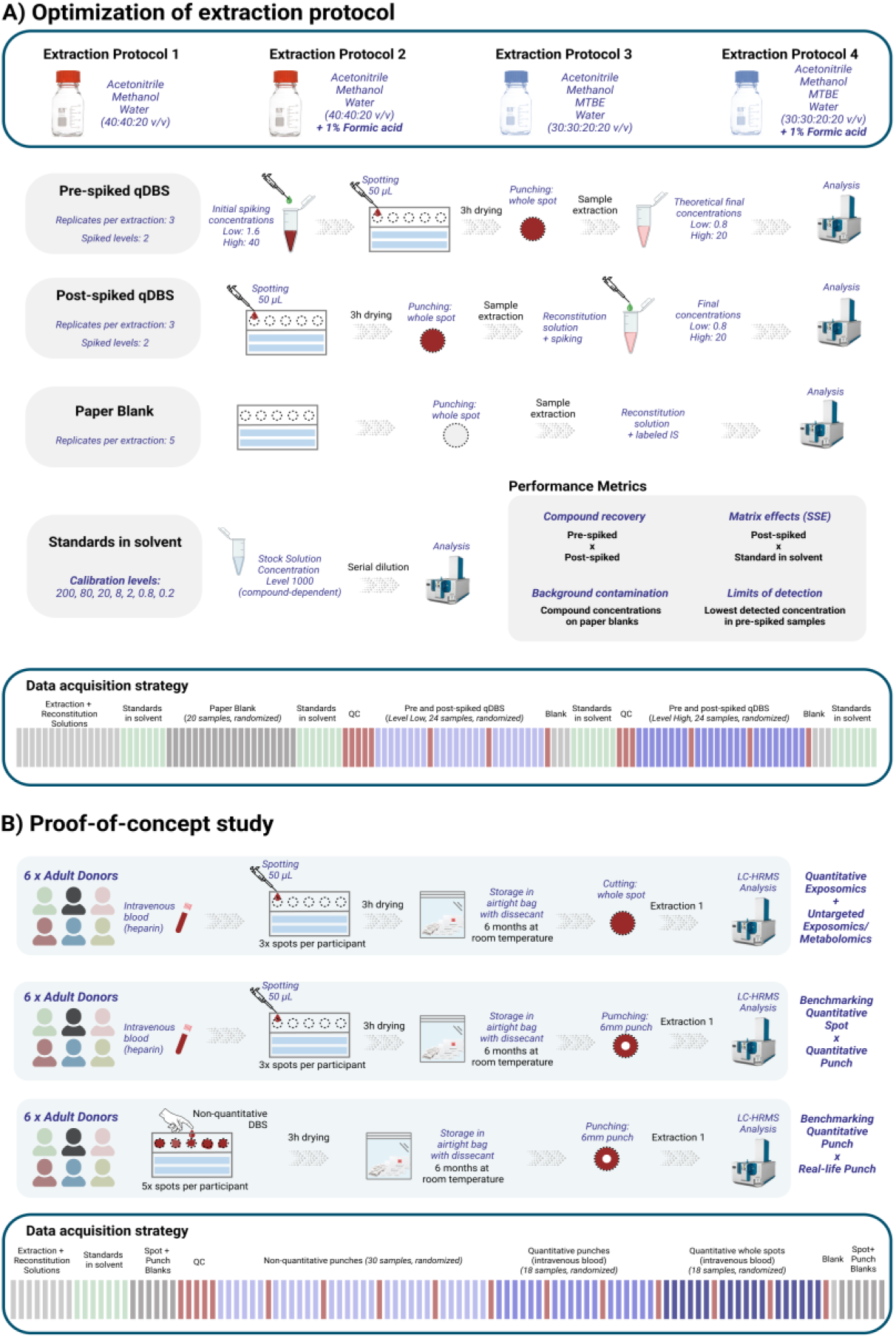
Experimental design and data acquisition strategy. After selection of best extraction protocol (Figure 1A), a proof-of-concept study using real-life samples obtained from six adult donors was performed (Figure 1B).

### 2.3. Generation of quantitative DBS samples (qDBS)

Spiked and non-spiked quantitative DBS (qDBS) samples were prepared on the day of blood collection employing the reverse pipetting technique. For non-spiked qDBS, 10 spots were produced per participant by pipetting 50 µL of heparinized blood into Whatman 903 Protein Saver cards. These samples were used for the proof-of-principle study and the targeted evaluation of xenobiotics as well as for a comparison with real-life dried blood spots collected as described in the previous section and further described in section 3.2.3. and, similarly to the samples described in the previous section, were also stored at room temperature for six months. Additionally, pre-spiked qDBS were prepared from participant #3 at ‘low’ and ‘high’ concentration levels. For that purpose, 75 µL of a stock mixture containing the >200 analytical standards (previously described in section 2.1) and labelled internal standards in acetonitrile were added to 1425 µL of heparinized blood, in compliance with a maximum 5% v/v ratio of blood to organic solvent, recommended to avoid protein precipitation and considerable disruption of the system^46^. Additionally, spiked blood was gently agitated for 30 min prior to be spotted onto the DBS cards. This time is recommended to allow for the partitioning equilibrium between red blood cells and plasma to be reached, better reflecting an *in vivo* exposure scenario^46,47^. Finally, post-spiked samples were generated by extracting non-spiked DBS and utilizing a spiked reconstitution solution in the next step in other to obtain post-spiked samples at the same concentration levels (‘low’ and ‘high’). All qDBS produced (spiked and non-spiked) were put to dryness for at least 3h in a ventilated space protected from direct light before extraction. In total, 223 exposure compounds with highly diverse chemical properties were included in the mixture (see **Table S1** for the detailed compound concentrations). The nominal concentration levels in the final extracts considering full analyte recovery and known analyte loss during supernatant transferring (**see section 2.5**) were at level 0.8 and level 20 for the ‘low’ and ‘high’ levels, respectively, as previously described in section 2.1. For assessing the impact of the haematocrit level on recovery and matrix effects, both pre– and post-spiked samples were employed, with a minor modification for pre-spiked samples from the previously described procedure. For this initial comparison, non-spiked qDBS from participants #5 and #1 were selected as the samples with the lowest (40.2%) and highest (48.9%) haematocrit level available, respectively. Next, the spiking solution was added to the selected qDBS rather than spiking the blood previously to spotting. Samples were spiked to obtain the theoretical level ‘high’ after extraction. Similarly, post-spiked samples were prepared at the same concentration level.

### 2.4. DBS cutting/punching

For the comparison of extraction performance, the whole spot was extracted by cutting the circle using an ordinary paper puncher with a slightly larger diameter than the blood spots (DBS diameter 12.7 mm, paper puncher diameter 16 mm). For the analysis of real-life DBS samples, experiments were performed from whole spots as well as 6 mm DBS punches, obtained via a pneumatic 6 mm DBS card puncher from Analytical Sales & Services Inc. (Flanders, NJ) for better reproducibility. In both cases, the devices were cleaned with a dry tissue between punching the same sample type/participant to avoid a possible particulate carry over. When switching between different sample types, devices were thoroughly cleaned with LC-MS grade acetonitrile and dried using pressurized air.

### 2.5. Sample extraction

Each DBS circle was inserted to a 1.5 mL plastic microtube followed by the addition of 1 mL of the different extraction solutions (**Figure 1**). The four solutions tested were named Ext#1-4 and solvent compositions were acetonitrile/methanol/water (40:40:20), acetonitrile/methanol/water (40:40:20) + 1% formic acid, acetonitrile/methanol/MTBE/water (30:30:20:20) and acetonitrile/methanol/MTBE/water (30:30:20:20) + 1% formic acid, respectively. Samples were mixed on a ThemoMixer (Eppendorf) at 1400 rpm for 5 min at 25°C, followed by a 10 min sonication in ultrasonic bath. Samples were then mixed for additional 5 min using the same conditions. After centrifugation at 10000 rpm for 5 min at 4°C, 800 µL of the supernatant was transferred to a clean microtube and taken to a concentrator at 20°C until complete solvent evaporation (overnight). Dried extracts were reconstituted with 80 µL of water/acetonitrile (90:10 v/v) and mixed for 10 min at 1400 rpm and 25°C on a ThemoMixer (Eppendorf). After centrifugation at 10000 rpm and 4°C for 5 min, the supernatant (60 µL) was transferred to an amber glass vial equipped with a 200 µL insert for LC-MS analysis. For the proof-of-principle study (**Figure 1**), extraction solution 1 was used following the same extraction procedure.

### 2.6. LC-HRMS data acquisition

A previously established LC-HRMS workflow for combined exposomics and metabolomics^42^ was employed for data acquisition. In brief, a 1290 Infinity II LC system (Agilent) coupled with a SCIEX 7600 ZenoTOF mass spectrometer was used. Chromatographic separation was achieved by reversed-phase separation (Acquity HSS T3 Column) over a 20-min gradient. Detailed parameters of the method are described in **Table S3**.

Data was acquired at both, MS^1^ and MS^2^ level, making use of either data-dependent (DDA) or data-independent (DIA, commercially named SWATH as sequential window acquisition of all theoretical mass spectra) acquisition modes to maximize MS^2^ spectra generation and improve metabolite annotation. The acquisition batches included the injection of pooled quality control samples (QC, see section 2.7) as well as an in-batch system calibration performed after each injection block (6 – 10 samples). First, an investigation on the most suitable extraction protocol among the 4 different extraction solutions was conducted using both spiked and non-spiked qDBS to produce pre and post-spiked sample extracts, respectively, with data acquired in DDA mode using an inclusion list for all 223 compounds. In this case, SWATH data was acquired for QC samples only. This approach was taken in order to ensure optimal fragmentation data for the compounds of interest with a potential increase in chemical coverage with SWATH data. For the proof-of-principle study, SWATH data was chosen as acquisition mode for the study samples while DDA data was obtained from QC samples. This approach was taken in order to maximize the coverage of fragmentation spectra obtained for (virtually all) detected ions in the samples, which might present compounds/ions specifically detected for each single individual. At the same time, acquiring DDA data on QC samples only allows for the acquisition of high-quality fragmentation spectra for representative and mostly abundant ions. The advantages of such data acquisition strategy were previously described by Guo and co-authors^48^. The detailed experiment design and data acquisition strategy is summarized in (**Figure 1**).

### 2.7. Quality control of analytical performance

Non-spiked qDBS pooled after extraction were used as quality control for both the evaluation of extraction performance and the proof-of-principle study. For the first, QC samples were prepared by pooling four non-spiked qDBS, where each sample was extracted using one of the four extraction protocols and pooled after reconstitution. For the proof-of-concept study, one additional qDBS was prepared for each participant (n=6) and QC samples were prepared by pooling all samples after reconstitution. Pooled QC samples were used to assess overall variability (relative standard deviation, RSD) of both analytical runs using three complementary approaches. First, RSD was calculated for labelled internal standards included in the reconstitution solution, for both ESI+ and ESI-. Results indicated an RSD <15% for labelled internal standards in all cases. Secondly, RSDs were determined for 8 compounds from the targeted list of xenobiotics that were found to be detected in the pooled QC sample. Finally, a summary on the variability of both unknowns and annotated features is also presented (see section 2.8 for more information on this topic). A complete description of the labelled internal standards included as well as a detailed summary of all quality control metrics are provided in **Table S4**.

As background contamination is typically observed for a handful of analytes included in this study, three types of blank samples were included in all the analytical runs to control for this issue. First, each extraction solution was directly injected four times, with the first injection considered as system equilibration and not included in the data evaluation (i.e., n = 3 per extraction type). Next, the reconstitution solution was directly injected into the LC-MS system using the same strategy (four injections, last three included in the data evaluation). Finally, blank paper spots (n=5) were submitted to the previously described sample extraction protocols and also included in order to evaluate the contribution originating from the paper and the overall sample preparation protocols.

### 2.8. Data analysis

Data processing was carried out using a two-step approach with (1) (semi-)targeted and (2) non-targeted data evaluation. Peak areas for all 223 xenobiotics of interest were determined using SCIEX OS software via extracted ion chromatograms based on *m/z* and expected retention time. Extraction window was set at 0.02 Da as recommended by the vendor. As an additional check, DDA data was inspected, especially when isoforms were to be expected. For peak integration, MQ4 algorithm was employed with general recommended settings, including For a few compounds, these parameters were slightly modified as needed to achieve reliable peak integration after visual inspection. This was preferred over manual peak integration to avoid operator bias. Nevertheless, for a handful of cases, manual peak integration was performed.

Peak areas were used to calculate analyte recovery and matrix effects (SSE). Recovery was defined as the ratio of average peak areas (n=3) from samples spiked before extraction to the average peak areas (n=3) from samples spiked after extraction. Since QC samples were the only non-spiked samples included in this experiment, peak areas were not used to subtract from pre or post-spiked before calculating recovery since QC samples were a pooled of all four extraction mixtures tested. Therefore, for compounds reported as detected in QC samples (see previous section), recovery values reported should be carefully considered. Instead, blank paper spot samples were used for this purpose (n=5)

After the selection of the most suitable extraction solvent mixture (based on recovery, SSE and practical aspects), an ‘order-of-magnitude estimation’ was determined for the method’s limits of detection (LOD), based on the samples spiked before extraction at both levels (‘low’ and ‘high’). In summary, compounds were assigned to 10-fold concentration ranges based on three possible scenarios. For the cases in which a peak was observed only for level ‘high’, there was enough evidence to define LOD at some point between level ‘low’ and ‘high’. For the cases in which a peak was not detected for neither level ‘low’ nor ‘high’, we corrected the lowest detection concentration in solvent for recovery and matrix effects to estimate LOD range. Finally, if a peak was observed at both levels, LODs were clearly around and/or below level ‘low’. In these cases, peak heights were used to assign LOD ranges. Since the data generated by this instrument typically uses a threshold of 100 cps for defining a detected peak, we have set a threshold of peak height at 1000 cps (as a less borderline detected peak) to define on the LOD range. Compounds detected below this value were assigned to the range for concentration level ‘low’. Otherwise (peak height > 1000), compounds were assigned to a LOD range immediately before the concentration of level ‘low’ (i.e., 10x lower in concentration).

It is important to highlight that this approach is rather conservative, i.e., the reported limits of detection are likely to be overestimated. Despite the relatively broad range reported for LOD values (10-fold), this approach was preferred since it considered a more realistic estimation than typical S/N and/or standard deviation-based calculations, usually recommended for low-resolution MRM-based quantification. From the authors’ perspective, both approaches would ultimately yield inaccurate/unrealistic values due to the limited number of replicates available for standard deviation calculations as well as the constraints associated with estimating S/N for high-resolution data. Therefore, presenting a comparatively large range instead of a single value provides a more pragmatic and realistic assessment to be used as reference values in HRMS studies of DBS in the future.

For obtaining preliminary data on the impact of the haematocrit levels in the reported quantitative data, peak areas for pre-spiked, (PRSDBS, in triplicate) and post-spiked qDBS (PTSDBS, in triplicate) were compared between the participants with the lowest and the highest haematocrit levels available, at 40.2 and 48.9%, respectively using independent Welche’s t-test performed in Metaboanalyst. Raw peak areas were used for PTSDBS samples while matrix effects-corrected peak areas were used for PRSDBS. This approach was chosen in order to maintain the original data variability of each replicate. In the case of PTSDBS, no correction is needed since the representation of matrix effects should be directly linked. In the case of PRSDBS, correction for matrix effects is needed to detangle this factor from compound recovery per se. For significance, we have considered FDR corrected p-value < 0.1. This value was selected instead of the typical 0.05 threshold due to the low number of replicates. Additionally, we assessed recoveries and matrix effects using an additionally analysed compound mix in solvent at the same nominal concentration. All results are displayed in Table S10.

Non-targeted analysis was evaluated by MS-DIAL (version 5.4.241021). Peak detection followed by alignment, deconvolution (SWATH data only) and annotation were performed. All parameters are described in details in **Table S5.** For metabolite annotation, Mass Bank of North America, MS-Dial and European Mass Bank databases were used, attributing confidence levels based on Schymanksi *et al*^49^, with a rather conservative approach. Initially, filtering for minimum dot product, weighted dot product and reversed dot product (400, 400 and 600) was used, followed by visual inspection of MS/MS matches. A maximum of 5 ppm and a minimum of two matching fragments was used for reporting a compound. Compounds with at least three matching fragments, no additional peak with the same match and no isomers included in the listed matching compounds were attributed level 2a. Compounds with at least three matching fragments, with either additional peaks (neighbouring or at different RT) with similar match pattern and score or with isomers (mostly positional) listed as matching compounds with similar match pattern were attributed level 2b. A more detailed description for such cases is provided in **Table S11**. Finally, compounds with only 2 matching ions were attributed level 3. Matches with non-specific fragments only (such as H2O or CO2 loss) were not included as well as annotations at level 4 or 5. The complete list of annotated compounds is available in **Table S11**. Matching scores and additional described parameters were described using a reference sample. Whenever possible, preference was given to fragmentation spectra acquired via DDA (QC samples as reference) over DIA data as spectra purity is expected to be higher.

## 3. Results and Discussion

### 3.1. Evaluation of extraction solvent mixtures

#### 3.1.1. Background contamination in filter paper

Background levels were evaluated for all 223 compounds in extracted blank paper spots as well as in the solutions used for sample extraction and reconstitution. This information is particularly important for exposomics studies once many of the targeted environmental contaminants are often widespread, including in house and laboratory dust, labware and, many times, LC-MS system parts. In the specific case of DBS samples, possible background contamination originating from the paper substrate might occur mostly via suboptimal storage conditions and/or inappropriate manipulation of the cards (e.g., without the use of gloves).

In total, 16 compounds were present in at least one of the three blank groups. For the majority of compounds, concentrations were found to be very similar among all the tested extraction conditions for both the extraction solution and the paper blanks. Exceptions were found mostly for extraction protocol #4, which presented higher concentrations of 2 –naphtol, 2-phenylphenol and atrazine. **Table 1** describes the compound distribution across all blank types for extraction protocol #1 (as the later method of choice) as well as the possible origin of contamination. Detailed data including quantitation parameters, extracted ion chromatograms and MS/MS spectra can be found in **Table S6.**

**Table 1.** Detected background contaminants over different types of blank samples.

| Compound | Background levels | Remark |
| --- | --- | --- |
| 2-naphthol | Extraction solution: N.D.<br>Reconstitution solution: N.D.<br>Paper blank: 0.1 ng/mL | Marker for naphthalene exposure, a polyaromatic hydrocarbon classified as industrial pollutant and probably carcinogenic to humans. Possible origins are as a residue from the paper card manufacturing process and/or via air contamination.<sup>50</sup> |
| 2-phenylphenol | Extraction solution: N.D.<br>Reconstitution solution: N.D.<br>Paper blank: 0.6 ng/mL | Potential endocrine disruptor extensively used in agriculture and industry and widespread pollutant in aquatic environments. Possible contamination from manufacturing process. |
| 4-methylbenzophenone | Reconstitution solution: N.D.<br>Extraction solution: N.D.<br>Paper blank: 23 ng/mL | Photo-initiator in packaging and ingredient for UV filter creams. Possible contamination from packaging or card handling without gloves.<sup>51,52</sup> |
| Atrazine | Reconstitution solution: N.D.<br>Extraction solution: N.D.<br>Paper blank: 0.09 ng/mL | Widespread herbicide in agriculture known for its persistence in soil and water as well as possible contamination to plant-based products<sup>53</sup>. Detection seem to be from paper substrate and might originate as a side contamination in the laboratory |
| Bisphenol A | Reconstitution solution: 0.13 ng/mL<br>Extraction solution: 0.18 ng/mL<br>Paper blank: 0.3 ng/mL | Plastics-related chemicals with xenoestrogenic activity. Overall concentrations indicate a close-to-zero contribution from the paper card itself since average concentration roughly represents the sum of both the reconstitution and extraction solutions concentrations. |
| Bisphenol F | Reconstitution solution: N.D.<br>Extraction solution: 0.05 ng/mL<br>Paper blank: 0.05 ng/mL |  |
| Dibutyl/<br>Diisobutyl phthalate | Reconstitution solution: 14.1 ng/mL<br>Extraction solution: 20.4 ng/mL<br>Paper blank: 16.1 ng/mL | Systematic contamination as represented by higher concentration in extraction solution. Previously reported as labware contamination<sup>54</sup> and indoor air of laboratories<sup>55</sup> |
| Methylparaben | Reconstitution solution: N.D.<br>Extraction solution: N.D.<br>Paper blank: 0.14 ng/mL | Preservative commonly used in cosmetics and pharmaceuticals for its antimicrobial properties. Possible contamination from handling the cards. |
| n-butylbenzenesulfonamide | Reconstitution solution: 0.6<br>Extraction solution: 1.4<br>Paper blank: 1.5 ng/mL | Plasticizer and emerging toxicant with potential neurotoxic effects<sup>56</sup>. Previously reported in house dust<sup>57</sup> |
| Nonylphenol | Reconstitution solution: N.D.<br>Extraction solution: N.D.<br>Paper blank: 25.1 ng/mL | Widely used compound for the production of various daily consumer goods<sup>58</sup>, with possible contamination via of the paper substrate via air or as a production side product. |
| Perfluorobutanesulfonic acid | Reconstitution solution: N.D.<br>Extraction solution: N.D.<br>Paper blank: 0.08 ng/mL | One of the typical PFAS reported before as background contamination in DBS paper substrates<sup>59</sup> |
| p-nitrophenol | Reconstitution solution: N.D.<br>Extraction solution: N.D.<br>Paper blank: 0.1 ng/mL | Common environmental contaminant with multiple sources, including diesel exhaust, textile industry and pesticide use. Contamination via air or card manufacturing is possible. |
| Tri-n-butyl phosphate | Reconstitution solution: 0.44 ng/mL<br>Extraction solution: 1.7 ng/mL<br>Paper blank: 1.6 ng/mL | Organophosphate esters, widely used as flame retardants and plasticizers. Evidence suggests endocrine-disrupting behaviour<sup>60</sup> and neurotoxicity<sup>61</sup>, including on prenatal exposure context<sup>62</sup>. Previously reported as labware background contamination<sup>54</sup> |
| Triphenyl phosphate | Reconstitution solution: 0.23 ng/mL<br>Extraction solution: 0.8 ng/mL<br>Paper blank: 8.9 ng/mL |  |
| Tris(2-chlorethyl)phosphate | Reconstitution solution: N.D.<br>Extraction solution: N.D.<br>Paper blank: 0.5 ng/mL |  |

No PFOS or PFOA was detected as background contamination in the filter paper while previous papers^37,63^ have reported background ranges of 0.01 – 0.06 and 0.9 – 1.1 ng/mL for PFOS and PFOA, respectively (ranges include reagent blanks, paper blanks and field paper blanks). Considering that the lowest detected concentration for standard in solution is reported here at 1.2 ng/mL, method’s sensitivity might be the reason behind this finding. Additionally, Poothong^64^ reported trace levels of perfluoroundecanoic acid (PFUnDA) and no detection of perfluorobutanesulfonic acid (PFBS), while the opposite is reported in this assay. For PFBS, an average background contamination of 0.03 ng/mL was found, a 10-fold lower concentration than reported by Lin^59^.

Alkyl phosphates such as tri-n-butylphosphate and tryphenil phosphate and tris(2-chlorethyl)phosphate were detected at particularly high concentration in all tested extraction solutions as well as in the final reconstitution solution, additionally to being detected in the extracted paper blanks. All compounds mentioned above (with additional alkyl phosphates) have been recently reported^54^ as common instrumental and laboratory background along with additional compounds here reported in paper blanks at similar concentrations of the reconstitution and/or extraction solution (namely di-(iso)butylphthalate and n-butylbenzenesulfonamide), reinforcing the assumption of a system contamination rather than originating from the paper substrate.

#### 3.1.2. Analyte recovery

Extraction recoveries were assessed for roughly 150 compounds at concentration level ‘high’ and 70 compounds at concentration level ‘low’. For the remaining compounds, recoveries could not be determined mostly due to non-detected peaks for both the pre and post-spiked matrix samples, with only a few cases for which post-spiked peaks were present. These results point towards a chromatographic and/or sensitivity limitation rather than the extraction process itself, based on the fact that even post-spiked samples did not present a peak. **Figure 2a** depicts the median and RSD values for all tested extraction conditions at level ‘high’, including the total number of compounds found to be within an acceptable recovery range (60 – 140%). Despite presenting a similar median value, protocols #2 and #4 were characterized by a higher data variability. A detailed description of the results is presented in **Table S7**.

**Figure 2.**
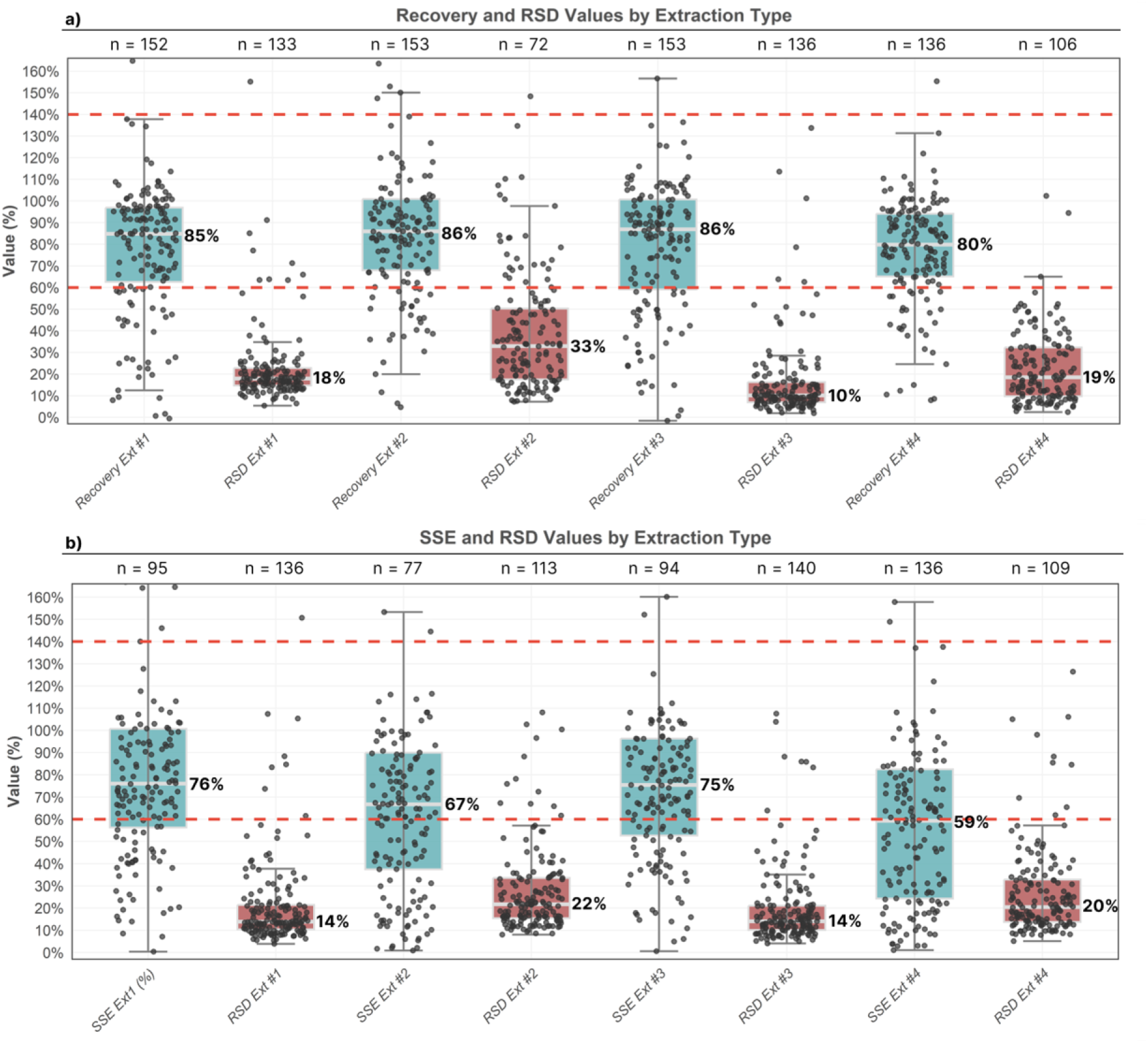
Compound recoveries (a) and matrix effects (SSE, panel b) are presented as boxplots (turquoise) alongside with RSD calculated based on technical replicates (n=3, red boxplots). Data is depicted for pre-/post-spiked samples at concentration level “high”. For every boxplot presented, median values (for either compound recovery, SSE or RSD) are also depicted on the right side of the boxplot. For visualization purposes, the y axis is limited to 160%. The horizontal dashed lines in red represent the interval for recovery/SSE between 60 and 140%. The displayed “n” represents either the number of compounds found to have recovery or SSE within the previously mentioned range (60-140%, turquoise boxplots) or the total number of compounds with RSD< 30% (red boxplots).

When comparing recoveries obtained at level ‘low’ and ‘high’ for compounds which recovery at level ‘low’ could be calculated, 40 out of 69 (58%) compounds were found to have a difference <20% and 47/69 (68%) <30%, demonstrating a fair agreement between concentrations that differ by a factor of 25. In addition, 75% of these compounds (51/69) presented higher recovery values at level ‘low’. Additionally, the comparison at level ‘high’ against validated recoveries described using the same extraction solution (extraction #1) for serum revealed 41 out of 49 (84%) compounds with differences in recovery below 30%.

Focusing on specific compounds, recoveries were recently reported^65^ for azithromycin in DBS as 91.9% at low level (3 ng/mL, 3 replicates) and 114% at level medium (500 ng/mL) for an optimized targeted method. Comparable recoveries were found at 112% (level ‘low’, 7.2 ng/mL) and 78% (level ‘high’, 180 ng/mL). For PFAS, recoveries were reported^59^ for a recent methodology employing bead homogenization followed by sonication and LC-HRMS analysis as 143 ± 12% for PFOA, 372 ± 5% for PFOS, 266 ± 23% for PFBS, all at 0.5 ng/mL, against 91 ± 20%, 95 ± 12% and 102 ± 17% reported here, respectively, at 0.3 ng/mL. For PFOS and PFOA, these values are comparable to the findings of Ma (83 ± 2% and 95 ± 4%) and Poothong^64^ (105 ± 18% and 95 ± 10%), respectively, at similar concentration levels. In the same study, recovery for BPA was described as 41 ± 4% (at 5 ng/mL) against 71 ± 18% described here at lower concentration level (2 ng/mL). For mycotoxins, Osteresch^39^ reported recovery values for an optimized targeted assay for quantifying 27 mycotoxins in dried blood spots. A direct comparison between our findings and reported values, respectively and considering similar concentration levels can be depicted for aflatoxins B1 (141 ± 20% x 100 ± 4%), B2 (93 ± 6% x 113 ± 10%), G1 (43 ± 7% x 81 ± 3%), G2 (65 ± 9% x 93 ± 3%) and M1 (56 ± 7% x 101 ± 4%), alternariol (68 ± 22% x 88 ± 4%), alternariol monomethyl ether (87 ± 25% x 110 ± 2%), ochratoxin A (90 ± 14% x 108 ± 5%), ochratoxin alpha (114 ± 26% x 89 ± 5%), T-2 toxin (93 ± 19% x 114 ± 5%), zearalanone (85 ± 13% x 100 ± 4%) and zearalenone (82 ± 12% x 101 ± 3%). Despite comparable, the slightly better results reported previously are likely related to the inclusion of acetone in the extraction solution. Finally, DBS-based recoveries for methyl and propylparaben were previously reported to be around 75% for concentrations higher than 20 ng/mL, in contrast with 117 ± 16% and 80 ± 9% reported here, respectively. For additional compounds included in this study such as alkyl phosphates, phthalates and polyphenols, no reference values were found to be previously reported in DBS.

Overall, no major differences on recovery or matrix effects (see section 3.1.3.) were observed between the samples with minimum and maximum haematocrit in the preliminary experiment (Table S10). A total of 72% and 88% of the chemicals showed a difference <10% for recovery and matrix effects, respectively. This confirms that our method is fit-for-purpose for the majority of compounds.

#### 3.1.3. Matrix effects

Matrix effects were more dependent on the presence/absence of formic acid in the extraction solution then on the chemical composition of the extraction solution, i.e. matrix effects were more similar between Ext #1 and Ext #3 (both non-acidified) as well as between Ext #2 and Ext #4 (acidified at 1% formic acid), as noted by the similar signal suppression/enhancement (SSE) values depicted in **Figure 2b**. As observed by the median values and the lower RSDs, SSE effects were observed to be less pronounced for extractions #1 and #3. In addition, matrix effects were found to be comparable to previously published values obtained using the same extraction conditions for Ext #1 using both serum^43^ and urine^44^ as detailed in **Table S7**. Considering only the overlapping compounds for which SSE is described for the tested methodology and literature reports (56 for serum, 32 for urine), SSE was found to be marginally more pronounced, with median values of 73% (DBS) against 81% (serum). For urine instead, median SSE was observed at 112% while median DBS value was found at 93%. Based on these findings, we can infer that there might be an additional contribution to ion suppression from species extracted from both the red blood cells and the DBS cards, while this contribution appears to be mild, especially when compared to a matrix with high salt content such as urine. On the other hand, it was also possible to notice very similar SSE values for different compounds in both cases. For serum, 3-benzylidencamphor (20% x 17%), trans-hydroxycotinine (61% x 66%), benzophenone (75% x 79%) and beta-zearalenol (84% x 90%) are some examples for DBS and serum, respectively. In addition, chloramphenicol (91% x 97%), fenarimol (52% x 56%) and fluconazole (41% x 49%) and imidacloprid (85% x 92%) can be highlighted when comparing DBS and urine samples, respectively.

A few compounds have also been reported^65^ previously in LC-MS based methodologies using DBS. For azithromycin, SSE values of 98% and 93% were reported at low and medium levels (3 ng/mL and 500 ng/mL, respectively), against 95% and 50% for levels ‘low’ (7.2 ng/mL) and ‘high’ (180 ng/mL) here described. SSE values were also reported for mycotoxins^39^ based on ratio of the slopes between pure solvent and matrix-matched calibration curves. For a more realistic comparison, the middle point between SSE values (and RSDs) obtained for levels ‘low’ and ‘high’ is here depicted against the literature, respectively, for aflatoxins B1 (61 ± 47% x 51 ± 4%), B2 (75 ± 10% x 423 ± 17%), G1 (76 ± 11% x 49 ± 7%), G2 (70 ± 19% x 54 ± 10%) and M1 (70 ± 40% x 57 ± 10%), ochratoxin alpha (94 ± 9% x 11 ± 8%) and T-2 toxin (73 ± 18% x 35 ± 7%), showcasing comparable performance, with mostly lower SSE effects reported here (despite higher RSDs). For PFAS, Lin *et al*^59^ have reported SSE mean values at 139% for a set of 24 PFAS (individual SSE were not reported), including all 7 PFAS reported here at mean value of 199% at concentration ‘high’. As highlighted by those findings, signal enhancement was rather the trend for PFAS in this methodology, especially at ‘low’ level for which a peak was observed in matrix but not in pure solvent for PFOA and PFOS. The short chain parabens (namely methyl-, ethyl– and propyl-) have also presented a trend towards signal enhancement. At level ‘high’, matrix effects were 551 ± 17%, 140% ± 4% and 104 ± 8% for methyl, ethyl and propylparaben, respectively (with methylparaben likely influenced by the background contamination). At level ‘low’, a similar behaviour was observed since peaks were only detected in spiked matrix (not detected in pure solvent at the same concentration level). Yakkund *et al*^38^ previously reported SSE of 98 and 97% for methyl and propylparaben, respectively. This difference is likely related to the fact that SSE are here reported for concentrations in the range 0.1 – 1 ng/mL for these compounds (level ‘high), while a concentration of 20 ng/mL (LOQ) was previously used^38^. For BPA, SSE was found to be roughly 40% based on the comparison of pure solvent and matrix-matched calibration curve by Ma, while 74% is described here for level ‘high’.

#### 3.1.4. Limit of detection (LOD)

The method’s LOD values were estimated based on the detection of a peak at levels ‘low’ and/or ‘high’ for samples spiked before extraction. Therefore, the values represent the estimated range of concentration in blood that the method is capable to detect. For exposure compounds not detected in samples spiked before extraction (24 out of 223 compounds), the instrument LOD is described, i.e., defined as the lowest concentration from the calibration curve for which a peak is observed. A detailed description of the results is presented in **Table S8**.

Overall, the majority of compounds presented LODs in the range of 0.1 to 1 ng/mL (84 out of 223) with a similar number of compounds in the immediate upper (10 – 100 ng/mL, 41 compounds) and lower ranges (0.01 – 0.1 ng/mL, 37 compounds). More specifically, LODs in the range of 0.01 – 0.1 ng/mL were estimated for all eight aflatoxins included, which was similarly reported by Osteresch et al. (with exception of AFB1, reported at LOD 0.0059 ng/mL). For PFAS, a comparison with reported values from Lin^59^ revealed matching ranges, such as 0.01 – 0.1 ng/mL for PFOA (0.083 ng/mL), 0.1 – 1 for perfluorononanoic acid (0.299 ng/mL) and perfluorohexanoic acid (0.132 ng/mL) as well as a higher LOD for perfluorodecanoic acid (1 – 10 ng/mL), reported as not detected^59^. For PFOS and PFBS (0.001 – 0.01 ng/mL), higher LODs were previously described (0.09 and 0.027 ng/mL, respectively). Finally, it is important to highlight the fact that high-resolution MS is expected to underperform when compared to low-resolution instruments^66^. In other words, the provided extraction method would certainly provide lower LODs if analysed via targeted, MRM-based assays.

### 3.2. Proof-of-concept study

Based on the results obtained, Ext #1 and Ext #3 protocols were found to have very similar performances for the targeted compounds. For practical reasons such as the use of three solvents instead of four and the possible difficulties involved in the pipetting of highly volatile solvents such as MTBE, Ext #1 was selected as most suitable. To showcase the method capabilities for the detection of both exogenous and endogenous compounds, a proof-of-concept analysis was followed. It is important to highlight that the proof-of-principle study relied on the same sample type (50 µL quantitative DBS) as used for assessing recoveries, matrix effects and limits of detection during the extraction efficiency comparison on solvent mixtures and, therefore, similar performance should be expected. In other words, recoveries, matrix effects and limits of detection are likely to change if different volumes are used.

#### 3.2.1. Targeted detection of xenobiotics in qDBS

In total, 11 compounds could be detected (**Table S9**), while concentrations could be reasonably estimated for 8 compounds (**Figure 3**). For a compound to be reported as detected, calculated concentrations corrected for recovery and matrix effects would need to be at least 30% higher than calculated concentrations in the blank (when present). Additionally, as quantification accuracy is a common limitation in HRMS assays, quantification confidence levels according to the scheme recently proposed by Petrick and co-authors are reported (**Table S9**). To the best of our knowledge, we here described for the first time the detection of 2-naphtol, 2-phenylphenol, 4-methylbenzophenone, and atrazine in DBS samples, while cotinine^36^, daidzein^67^, genistein^67^, PFOA^59,64^, PFOS^59,64^, propylparaben^38^ and trans-3-hydroxicotinine have been previously reported. Diverse chemical classes/applications are therefore covered, including industrial pollutants (phenols, PFAS), pesticides (triazines), dietary markers (polyphenols), smoking markers (nicotine derivatives) and personal care product ingredients (parabens).

**Figure 3.**
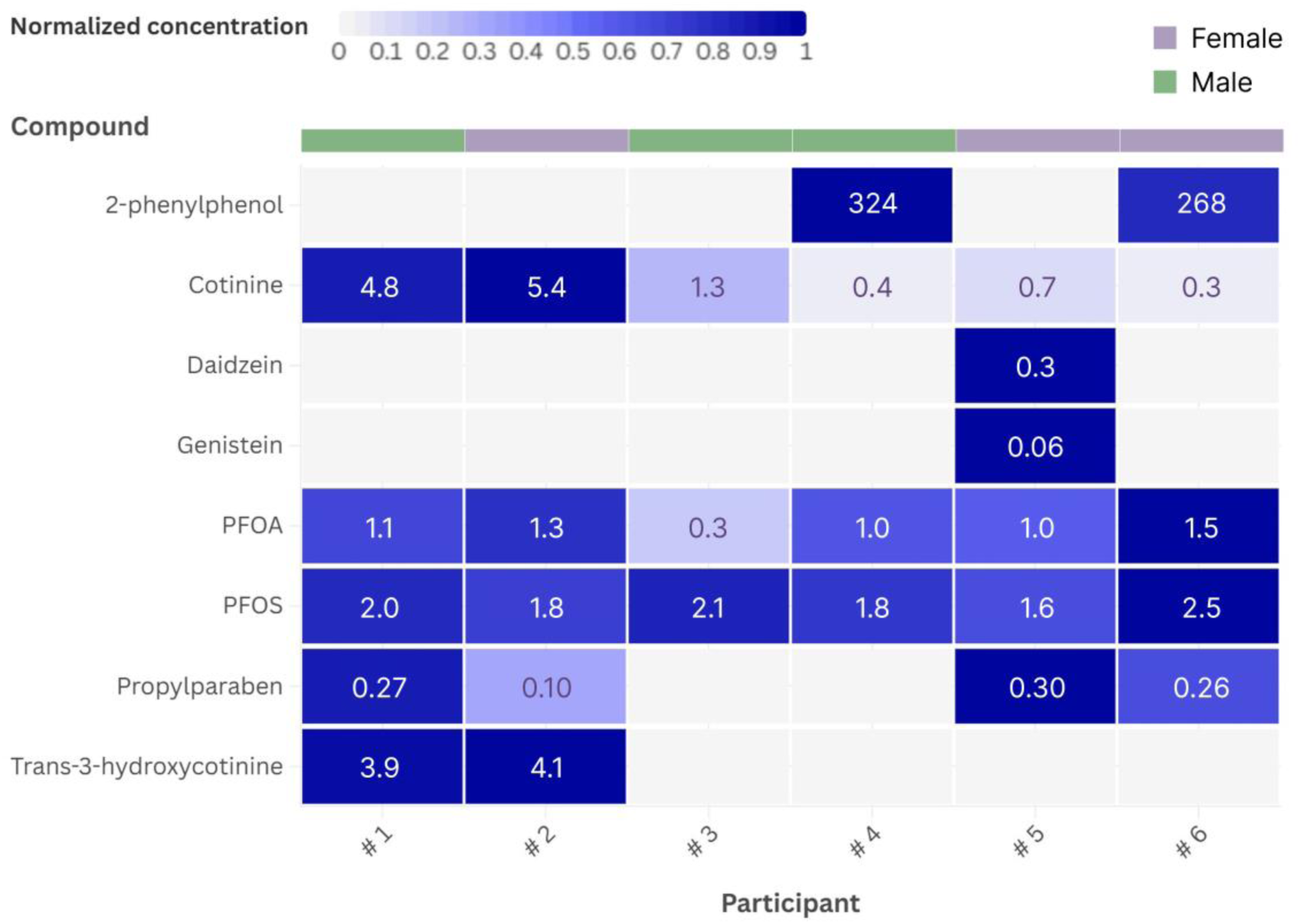
Individual exposome assessment from DBS samples from participants in the proof-of-principle study (3 male, 3 female). Colour scale reflects normalized concentrations per compound, with highest detected concentration established as 100%. Estimated concentrations are displayed in ng·mL^-1^. For some compounds, a more homogeneous exposure pattern is observed (e.g. PFOS) while highly participant-dependent exposure patterns are observed for others (e.g. cotinine, genistein and daidzein).

Yang *et al*^68^ have previously reported a method for quantifying cotinine in DBS of newborns as a marker for maternal smoking, defining concentrations higher than 6 ng/mL as the lower threshold for active smokers. Despite focusing on a different study population, our results are in line with this definition, once all participants (none being an active smoker) presented cotinine levels below this threshold. In addition, higher cotinine levels were observed for participants #1 (4.76 ± 2.82 ng/mL) and #2 (5.40 ± 3.19 ng/mL), who declared to be an occasional and a passive smoker, respectively. For participant #3, cotinine levels were about three to four times lower in concentration (1.29 ± 0.86 ng/mL), while a maximum of 0.74 ng/mL was found among the remaining participants (all non-smokers).

For PFOS and PFOA, despite the potential poor accuracy for calculated concentrations relying on two calibration points only, results are in accordance with previously reported concentrations. Ma *et al*^37^ used 192 DBS from New York State newborn screening program to detect and quantity PFOS and PFOA and PFOS with mean values of 1.28 ± 0.73 described for PFOA. More recently, Taibl et al^69^ quantified both compounds in 267 DBS measured in mothers during early/middle pregnancy, reporting mean values of 0.57 ± 2.31 for PFOA. Our results were highly in line with these findings, with mean value falling in between these reports at 1.06 ± 0.28 ng/mL). For PFOS, the same studies have reported mean values of 1.80 ± 0.98 ng/mL^37^ and 1.43 ± 2.7 ng/mL^69^. Similarly, we here report mean concentrations of 2.0 ± 0.3 ng/mL. Nevertheless, it is worth mentioning that the calculation of concentrations for PFOS was corrected for the SSE calculated for PFOA since this value could not be established for PFOS. This was done because no peak was observed in the solvent calibration samples for PFOS, while a peak was observed for the post-spiked DBS at the same concentration level (‘high’). This clearly indicates relevant signal enhancement; however, the quantitative estimation needs to be interpreted with caution. Since PFOA is not only the chemical most similar compound to PFOS but have also presented similar performance in terms of recovery at both levels ‘low’ and ‘high’, we have opted to proceed in this manner.

Daidzein and genistein also presented an interesting pattern, being detected in the only participant (#5) self-reported as vegetarian. Belonging to the same class of polyphenols (isoflavones), these compounds are mostly found in soybeans and soy products, with mean plasma concentrations found to be 23 times higher in vegans/vegetarians than omnivores in European adults.^70^

#### 3.2.2. Non-targeted exposomics/metabolomics

Aiming at increasing the described chemical space, non-targeted data analysis was performed for the annotation of exposure compounds not included in the semi-targeted investigation as well for the characterization of the accessible part of the metabolome. **Figure 4** presents normalized peak areas for all participants, illustrating a diverse set of endogenous metabolites, xenobiotics and diet-related compounds that could be confidentially annotated via MS/MS spectral matching against publicly available spectral libraries. The complete set of annotated compounds (levels 2a and 2b^49^) can be found in **Table S11**. It is worth to highlight the agreement between peak areas and expected testosterone levels, with similar peak areas found among participants from the same sex, with a large gap between males and females (higher in males). In addition, it is possible to observe that with the exception of testosterone and glycocholic acid, metabolite levels appear to be more homogenous across participants, while exposure compounds present a larger variability, demonstrating the individualized aspect of the exposome. Caffeine and piperine, for instance, are markers of very specific food consumption (coffee and black pepper) and, therefore, will highly be influenced by dietary habits. It is also worth noting the annotation of gabapentin, a drug used to treat neuropathies, fibromyalgia, acting as an anticonvulsant and neuropathic pain management medication. As such, there is limited prescription across the average population and, in addition, no participant has actively reported to actively take the drug. Despite that, gabapentin was detected in all participants, with satisfactory MS/MS match. Previous works have described similar observations. Chao^71^ reported gabapentin in a significant portion of analysed human placental samples (annotation level 2^49^), suggesting potential sources beyond direct drug intake. Suhre^72^ and co-authors compared self-reported medication intake with the correspondent detection in blood stored at the Qatar Biobank for 2807 individuals and while only 0.1% self-reported the use of gabapentin, the parent compound was detected in 0.8% of the samples.

**Fig 4.**
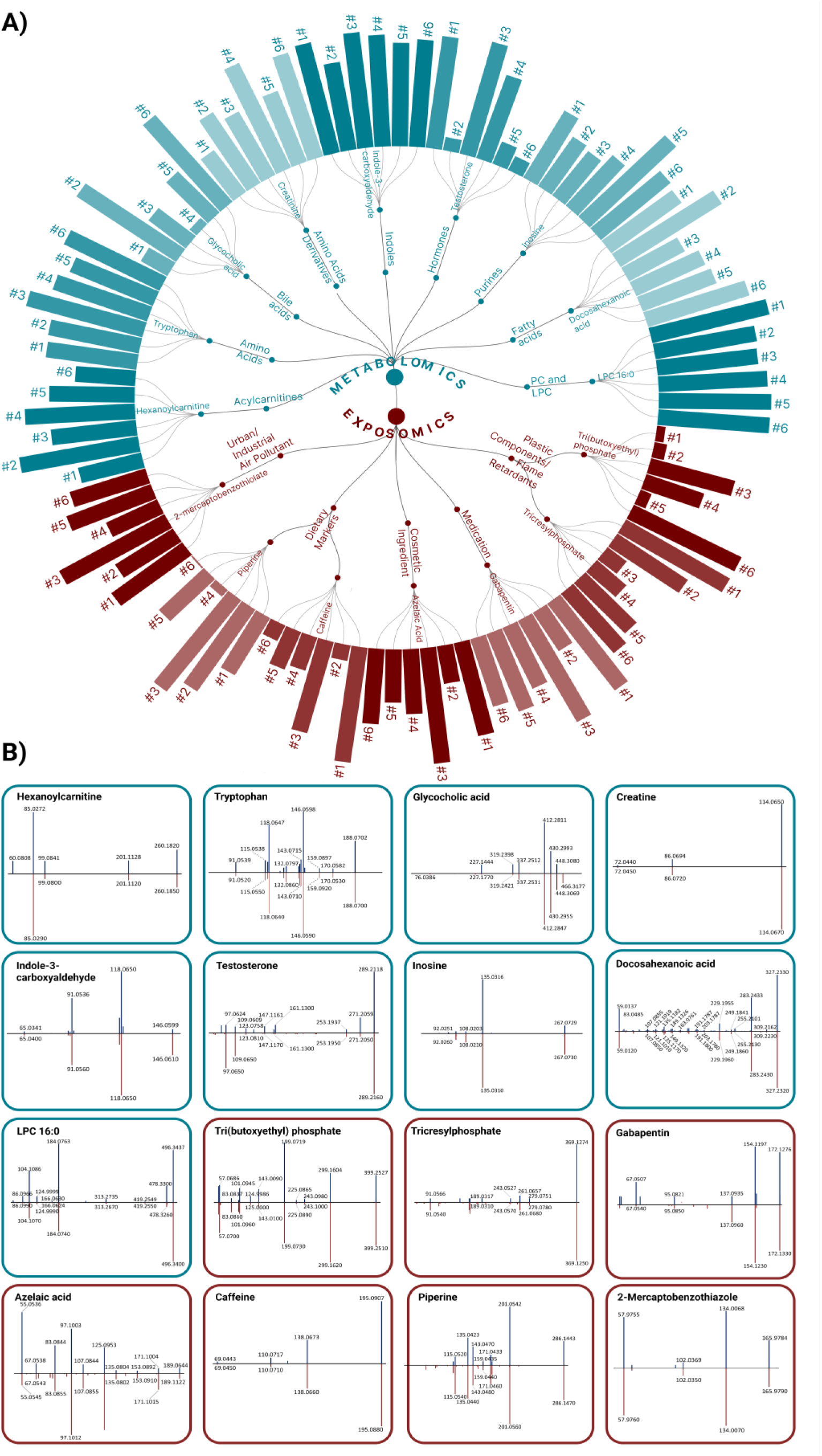
(**A**) Participant-dependent normalized peak areas for a diverse set of endogenous (turquoise and exogenous (red) compounds detected and annotated via non-targeted data analysis, alongside with correspondent MS/MS spectral match (panel B).

Recently, Thaitumu^73^ reported the annotation of 46 compounds (23 at annotation level 1 and 23 at annotation level 2) in a comparison among different microsampling devices (including DBS) via untargeted metabolomics. Sample extraction was similar to adopted in this work both in terms of extraction solution (methanol/water 60:40 v/v) and extraction techniques, combining shaking and sonication steps. Spectral matching also applied the same process in general, using MS-Dial against Mass Banks and MS-Dial libraries. From those, 36 (78%) were also successfully detected an annotated in our dataset (mostly at annotation level 2a)

#### 3.2.3. Exploratory analysis: whole spot x punch

In order to estimate the impact on the detection frequency of different xenobiotics, a comparison among three sample types was also included in the pilot study. In addition to the quantitative whole spots (SptQT, triplicate), 6 mm punches were obtained from qDBS (namely quantitative punch, PncQT) as well as from real-life DBS samples collected from the fingertip (namely non-quantitative punches, PncNQ). From all 15 compounds previously reported for the analysis of the whole spot, only 4 could be detected in PncQT or PncNQ: 2-naphtol, 4-methylbenzophenone, cotinine and trans-hydroxycotinine (2 participants only). Overall, calculated concentrations for PncQT and PncNQ presented a fair agreement for the majority of the cases. For 4-methylbezophenone, differences are evident (higher concentrations for PncNQ), which might be related to contamination of the card. PncQT were prepared in-lab with the use of gloves while PncNQ represent real on-field samples collected by the participants. When comparing to SptQT, PncNQ also showed a good agreement between expected and calculated concentration. Nevertheless, there a clear indication that the use of a single punch might limit the detection of a significant number of compounds. In addition, previous works^74,75^ have reported different concentrations for endogenous metabolites when comparing different punch locations (e.g. central x peripherical) and the same is likely possible for compounds of exposure. Therefore, despite the need of higher volumes for extractions – and consequently longer drying steps – the use of the whole spot seems to be more recommended to increase compound detection and sample representativeness. Results are detailed in **Table S12**.

## 4. Limitations

It is important to emphasize that the workflow can undergo further refinement. Along with the composition of the extraction solution, additional parameters could potentially influence recovery, SSE and LODs. Among many, different extraction approaches or longer extraction times are likely to be the two most influential factors and could be evaluated in the future. However, it is important to highlight that in the context of multi-class exposomics, especially employing HRMS, an optimal performance for all analytes is not feasible and not required, but rather a balance between sensitivity and chemical coverage should be achieved^5,76^. Additionally, the quantitative aspect can be further improved. It was possible to observe a large variability on detected concentrations across replicates, illustrated by a large RSD for some compounds/samples, especially for compounds with background contamination. As such, quantification confidence levels 4 and 5 are mostly reported. Strategies such as internal standard addition to the card prior to analysis and the use of a higher number of replicates can help in overcoming such limitation. Also related to that is the fact that all analytical indicators were obtained at a single haematocrit level whereas it is possible that differences may be observed when comparing samples with low and high haematocrit content, increasing quantitation bias. This is especially important and should be accounted for future applications focusing on quantitative data from newborns. Moreover, a systematic evaluation of compound stability was not performed in here whereas it might play an important role, especially if future applications of the method focusing on historical samples is foreseen. In addition, more analytes and chemical classes may be included to the targeted evaluation in the future. Finally, a large-scale application of the method in a newborn cohort is pending, with the proof-of-principle study being performed only on a limited number of samples.

## 5. Conclusion and Perspectives

The presented HRMS workflow highlights the potential of dried blood spots (DBS) as an alternative biospecimen for multi-class exposure assessment, coupled with a comprehensive metabolic profiling approach. Despite the extremely low concentrations typically expected for the environmental and food-related xenobiotics under investigation, which becomes even more challenging with such a sample type, it was possible to obtain satisfactory analytical performance (recovery, matrix effects, reproducibility and LOD), especially when considering the use of HRMS.

Based on the outcomes of this study, further investigation towards chemical exposure in neonates should be prioritized to obtain a baseline assessment on exposure levels. As a long-term perspective, the method might be further optimized for routine exposure analysis in newborn screening programs globally. Nevertheless, points should still be addressed in future studies, including a more comprehensive assessment on the impact of different haematocrit levels on compound concentrations. A few works have tried to also estimate this impact^77^ as well as to propose models for correction based on post-collection haematocrit^78^ and blood volume estimation^79^. Yet, from the authors perspective, high-accuracy full quantitation will likely remain a limitation related with such sample type. Future studies should address these challenges keeping in mind the fit-for-purpose criterion of any exposomics method. In the context of the omic-scale exposure monitoring in newborns, for instance, estimating relatively broad concentration ranges is likely enough to observe long-term trends and high-exposure scenarios potentially related to long-term health consequences. On a different note, scaling is also crucial and might include the use of 96-well plates during sample preparation as well as the use of automated systems for sample handling, enabling the fitness-for-purpose of the method for exposome wide association studies (ExWAS) in the future. Finally, a systematic evaluation of background contamination in real-life samples is needed. At this point, considering the suboptimal storage of most DBS samples for exposomics analysis purposes, we recommend a paired sample-blank analysis from the same card, from a point close to the collected blood droplet. The rationale behind this recommendation is the fact that, despite the manual handling of the cards is typical within newborn screening and/or biobanks, we speculate that operators tend to be less careful when touching the outer parts of the card when compared to the proximities of the designated spot for sample collection. Therefore, blank contamination might be overestimated and not representative if such outer locations are used as blank. As this might pose a limitation in terms of the high number of samples to be additionally analysed, aiming at running a representative number of blanks according to each study design should be pursued. Reaching all previously mentioned milestones will be key to achieve a biomonitoring status for such methodology with data accuracy and precision. In Austria alone, around 80,000 newborns are screened every year for metabolic diseases, with remaining collected samples being continuously stored, and centrally organized. As a particularly vulnerable population, monitoring possible exposure to chemicals, especially in high exposure scenarios, may be relevant for individual health prognosis. Large scale studies benefiting from these historical samples should be a priority to evaluate the findings and the feasibility of an exposomics monitoring approach to be consolidated as a routine screening in newborn programmes around the globe.

## Acknowledgments

The authors would like to acknowledge all members of the Warth lab for providing valuable discussions and Teresa Sopuch for technical support. The authors would also like to express their gratitude to the Mass Spectrometry Center of the Faculty of Chemistry, University of Vienna as well as to SCIEX Ltd. in the name of Antonella Chiapparino and Jean-Baptiste Vincendet for technical support. This work was supported by the University of Vienna via the Exposome Austria Research Infrastructure, the Austrian Federal Ministry of Education, Science and Research (project DigiOmics4AT), the Austrian Federal Minister of Innovation, Mobility, and Infrastructure and the Austrian Federal Ministry of Agriculture and Forestry, Climate and Environmental Protection, Regions and Water Management.

## Statement

During the preparation of this work the authors used ChatGTP in order to polish the language of minor sections. After using this tool/service, the authors reviewed and edited the content as needed and take full responsibility for the content of the published article.

## Data availability

Raw data files along with metadata are in the submission process into Metabolights (no repository code available yet). The files will be publicly available under the name “*Dried blood spots analysis for targeted and non-targeted exposomics*”

## Author contributions

**VVH**: Conceptualization, Methodology, Investigation, Writing, Reviewing; **MZ**: Conceptualization, Reviewing; **LW:** Conceptualization, Investigation, Reviewing; **BW:** Conceptualization, Methodology, Investigation, Writing, Reviewing, Supervision, Funding Acquisition

